# International Migrant Workers, Heat Exposure, and Climate Change: A Systematic Review of Health Risks and Protective Interventions

**DOI:** 10.1101/2025.06.04.25328218

**Authors:** Lara van der Horst, Oumnia Bouaddi, Sarah Williams, Meg Jago, Karen Lau, Ben Furber, Sally Hargreaves, Amirah Zafirah Zaini, Engy Mohamed El-Ghitany, Tharani Loganathan, Andreas Flouris, Davide Testa, Cathy Zimmerman, the Consortium for Migrant Worker Health

**Author notes:** Corresponding author Sally Hargreaves.

## Abstract

**Background:** International migrant workers, representing 170 million people globally, often face hazardous working conditions, including extreme heat exposure. This increases their risk of occupational heat strain, exacerbated by poor working conditions. This systematic review aims to identify the health risks of occupational heat exposure among international migrant workers globally, and document existing protective interventions and measures, in order to inform policies that protect this vulnerable population.

**Methods:** We searched four electronic databases (Medline, Embase, Ovid Global Health and PsychINFO) for primary research studies (January 2014–April 2024) on international migrant workers experiencing adverse health outcomes following high working temperatures. Records were screened, and data extracted by two independent reviewers. Assessment of study quality was done using Joanna-Briggs Institute checklists. Results were synthesised narratively and reported following PRISMA 2020 guidelines. The protocol was registered in PROSPERO (CRD42024519547).

**Results:** Of the 646 records screened, 19 studies involving 2,322 migrant workers across six countries were included in the analysis, with most studies from high-income countries (n=14, 74%), mainly the USA. Studies focused on workers in construction (48%) and agriculture (42%), with migrant workers originating from 14 countries, predominantly India, Mexico, and Nepal. Reported health outcomes included heat-related illnesses (n=12), dehydration (n=5), kidney disease (n=2), and poor skin health (n=2). Common symptoms included headaches, muscle cramps, and heavy sweating. Interventions focused on water, rest, shade, skin protection, and education, but evaluations were limited and some measures failed to address heat exposure effectively.

**Conclusions:** Occupational heat exposure poses significant health risks for international migrant workers. Where interventions exist, barriers to effectiveness remain, with a knowledge gap as to the situation in low- and middle-income countries. Amid rising global temperatures improved worker education, worker-tailored and co-designed interventions, updated protocols, and increased healthcare accessibility are urgently needed.

## Introduction

International migrant workers, defined as individuals who are to be engaged, are engaged or have been engaged in a remunerated activity in a State of which they are not nationals (1), make up 170 million of the world’s population and a significant portion of the labour workforce (2). The top recorded destinations of work being Europe, North America, and the Middle East, with millions of remittances received by low and middle income countries (LMICs) (2, 3). Migrant workers often work in irregular settings with limited legal protection making them more vulnerable to occupational health risks (4). Often, these workers are paid less than their non-migrant counterparts and are often subjected to worse working conditions that include more strenuous work, less flexible work schedules and exploitative financial arrangements (5–7). In addition, they are more likely to experience occupational health injuries and less likely to use health services than non-migrant workers (8). For instance, in a recent global meta-analysis among 7260 international migrant workers, the pooled prevalence of having at least one occupational morbidity was 47% (95% CI 29-64; I2=99·70%) (9). As a result of these trends, migrants are commonly referred to as working in 3D jobs, a term used to describe jobs that may be a combination of dirty, demeaning, demanding, dangerous, or difficult (10, 11).

Globally, it is estimated that 33.8% of international migrant workers are employed in farming, manufacturing, mining and quarrying, and construction (12). Workers in these sectors typically perform their duties outdoors, increasing their exposure to environmental heat. Especially when combined with heavy physical labour, such heat exposure can result in a range of adverse health effects collectively known as occupational heat strain (OHS). OHS can include dehydration, heat stroke, heat cramps, heat exhaustion and 15% of cases involve kidney disease or acute kidney injury (AKI) (13). Where legislation exists to protect workers, workers may still find themselves at risk. For example, the Gulf countries have implemented bans on midday working, and measures to ensure the availability of drinking water, shade, and PPE (14). However, an assessment of the ban on midday work has shown that it is not entirely effective and needs to be combined with the provision of shaded areas, access to cool water and rehydration salts, and medical checks (15). Similarly, a study among migrant workers in Kuwait found that they experience a three-fold higher risk of mortality from high temperatures (16). Worse heat-health risk profiles in migrant workers occur as a result of personal factors like a smaller body size, preferences for wearing more clothing, and working more intensely with fewer rest breaks (17). Furthermore, greater vulnerability to occupational heat strain also results from reduced access to resources for the prevention and treatment of heat-related illnesses which results from language and cultural barriers (18, 19).

There are multiple previous reviews exploring the effects of occupational heat exposure on workers’ health and productivity, finding that such exposure leads to OHS, more occupational injuries, increased risk of vector borne disease and reduced productivity (13, 18, 20–22). However, evidence is limited in terms of international migrant workers. El Khayat et al. identifies migrant workers as a vulnerable group calling for increased research into migrant workers specifically (18). Reviews investigating effects on migrant workers are fewer and confirm migrant’s vulnerability to occupational injuries (9), with another concluding that there is a high burden of heat-related illness amongst this group (23). This will be the first review to focus specifically on international migrant workers with an additional analysis on protective interventions to allow for necessary improvement in preventative efforts to protect this vulnerable group (23).

The United Nations (UN) Sustainable Development Goals has acknowledged the protection of migrant health as a global priority (24). The extension of the Global Action Plan Promoting the Health and Wellbeing of Refugees and Migrants, also demonstrates similar intention (25). The unique risk factors for occupational heat exposure among migrant workers, and their limited access to protective resources calls for strengthening health and labour policies targeted to this vulnerable group for these priorities to be met. Gaining a deeper understanding of the risks facing international migrants working in hot conditions is the first step to guiding the development of such policy.

## Methods

This systematic review is reported according to the Preferred Reporting items for Systematic Review and Meta-Analysis (PRISMA) 2020 guidelines (26). The protocol for this review was registered with PROSPERO, number: CRD42024519547.

### Aims

The primary objective of this research is to systematically identify the direct and indirect health risks in migrant workers following occupational heat exposure. The secondary objective of this research is to identify interventions and strategies that are being implemented to mitigate adverse health effects following occupational heat exposure in migrant workers. This systematic review will improve understanding of the vulnerabilities of migrant workers to occupational heat exposure, to inform global and national policy in migrant-receiving countries.

### Search strategy

We searched four electronic databases including Medline (Ovid), Global Health (Ovid), PsychINFO (Ovid), Embase (Ovid) and the Cochrane library for studies reporting health outcomes in migrant workers following occupational heat exposure published between 01/01/2014 and 31/04/2024. This period was selected to represent a time within which high environmental temperatures could be easily attributed to anthropogenic climate change. It aligns with the publication of the International Labour Organization’s *Guidelines for a Just Transition Towards Environmentally Sustainable Economies and Societies for All* (27), which represents an important policy landmark in identifying the need for occupational health and safety and social protection of workers in the face of climate change. Additionally, 2015 has been identified as an initial year in which labour productivity was significantly reduced due to the impact of heat stress on vulnerable workers (4). A search strategy combining terms for *heat*, *health*, and *migrant worker* was developed by L.V and refined with support from K.L. and university librarian. Previous reviews on similar topics were also used to inform the development of the strategy (9, 13). The full search strategy has been added to supplementary material table S1. In addition, we also searched key grey literature websites including the International Labour Organization (ILO), the International Organization for Migration (IOM), and The National Institute for Occupational Safety and Health (NIOSH). Relevant reports published by these institutes were searched for key references not picked up by the search strategy.

### Inclusion and exclusion criteria

Inclusion and exclusion criteria were developed using a population, exposure, and outcome framework, adapted from the population, intervention, comparison, outcome, and study design (PICOS) framework (28). The population included international migrant workers, defined as individuals who are or have been employed outside their country of origin. The exposure of interest was occupational heat. Due to the inconsistent use of heat exposure indicators (21) and the varying levels at which individuals may experience heat stress (29), heat exposure is often difficult to define, so this criterion was reflected in the search strategy. Outcomes of interest included poor health outcomes (defined as impaired health or well-being of the workers), and interventions aimed at mitigating the impact of occupational heat on the health of migrant workers. Any comparators were included (e.g. host populations, migrant workers non-exposed to occupational heat). We included all primary study designs, including cross-sectional, prospective cohort, and case-control studies, using both qualitative and quantitative approaches. Studies were excluded if they did not include migrant populations, if environmental temperatures made no contribution to high working temperatures, or if the study did not occur during working hours. Language was restricted to English, but no geographical restriction was applied.

### Study selection procedure

Records obtained from the searches were imported into EndNote to remove duplicates and then exported to the web-based application Rayyan (30), where remaining duplicates were removed. Titles and abstracts were screened by L.H; full-text articles included at this stage were retrieved and screened for eligibility by L.H; 25% of the screening process was duplicated by M.J. Disagreements at any stage were resolved through consensus.

### Data extraction

The information extracted included author, publication date, study design (e.g., observational, experimental), country of study, study setting (e.g., urban, rural), and period. Population characteristics included occupation, industry, country of origin, length of stay, age range, and gender. Exposure characteristics involved a measure or description of heat exposure. Interventions were defined as any action implemented by study participants, employers, or members of the research team aimed at mitigating poor health outcomes caused by exposure to high ambient temperatures. Information extracted about interventions included intervention characteristics (description of the intervention), positive outcomes, negative outcomes, and any documented impact. For studies with a majority migrant population, data referring to the whole population was used. For studies where migrants made up half or less of the population, only data regarding migrants was used. If studies had a comparator (e.g. host population), the same data was extracted from the comparator group.

### Quality assessment

We used the Joanna Briggs Institute (JBI) checklists (31), appropriate for the study designs of the included studies, to assess the risk of bias. A scoring system was used, where studies scoring between 0–3 were considered low quality, those scoring between 4–6 were considered of average quality, and those scoring between 7–9 were considered high quality, as was done in a similar review (9). Quality assessment was duplicated by M.J with disagreements resolved through consensus. Studies were not excluded based on quality.

### Data synthesis

The methodology of the results synthesis is that of narrative synthesis. The Economic and Social Research Council (ESRC) Methods Programme Guidance was used, paying particular attention to elements 2 and 3, *developing a preliminary synthesis* and *exploring relationships in the data*, respectively (32). Data were collated, then tabulated and displayed to best summarise the findings. The data were separated into subgroups where appropriate and compared accordingly. Qualitative data such as protective interventions were manually analysed for trends and common themes.

## Results

### Overview of included studies

Of the 646 records screened, 195 full-texts met our eligibility criteria, of which 19 studies involving 22 migrant workers across six countries were included in the final analysis (17, 33–50) (see Figure 1 – PRISMA flowchart). Studies were mainly conducted in high-income countries (HICs) (n=14, 74%); the United States (US) (n=13) and Bahrain (n=1) (34), with the remaining studies in low and middle income settings; Nepal (n=2) (39, 49), Costa Rica (n=1) (47), Cyprus (n=1) (17), and the Dominican Republic (n=1) (43). The studies in the USA were carried out in different sites including North Carolina (n=3) (37, 38, 44), Georgia (n=3) (40, 41, 45), Florida (n=1) (33), Iowa (n=1) (35), Mississippi (n=1) (48), Oregon (n=1) (42), South Carolina (n=1) (46), California (n=1) (36), and one unspecified (50) (see Figure 2). Study locations varied with three conducted at migrants’ places of work (17, 33, 35), ten in health care facilities (34, 39–41, 43, 44, 46, 48–50), five in worker accommodation or the community (37, 38, 42, 45, 47), and one unspecified (36). All studies reporting the work environment described all or some work occurring outdoors. Indoor work was limited and included agricultural work in packing sheds (45), 43% of construction workers who work partly or fully indoors (34), and workers in housekeeping, restaurant work and factory work in another study (41). The majority of studies had a cross-sectional design (N=9) (53–55, 58, 59, 61, 63, 65, 68). Nine studies were found to be of low quality (34, 37, 39, 42, 43, 45–47, 50), and ten of average quality (17, 33, 35, 36, 38, 40, 41, 44, 48, 49) (see Table 1).

**Figure 1:**
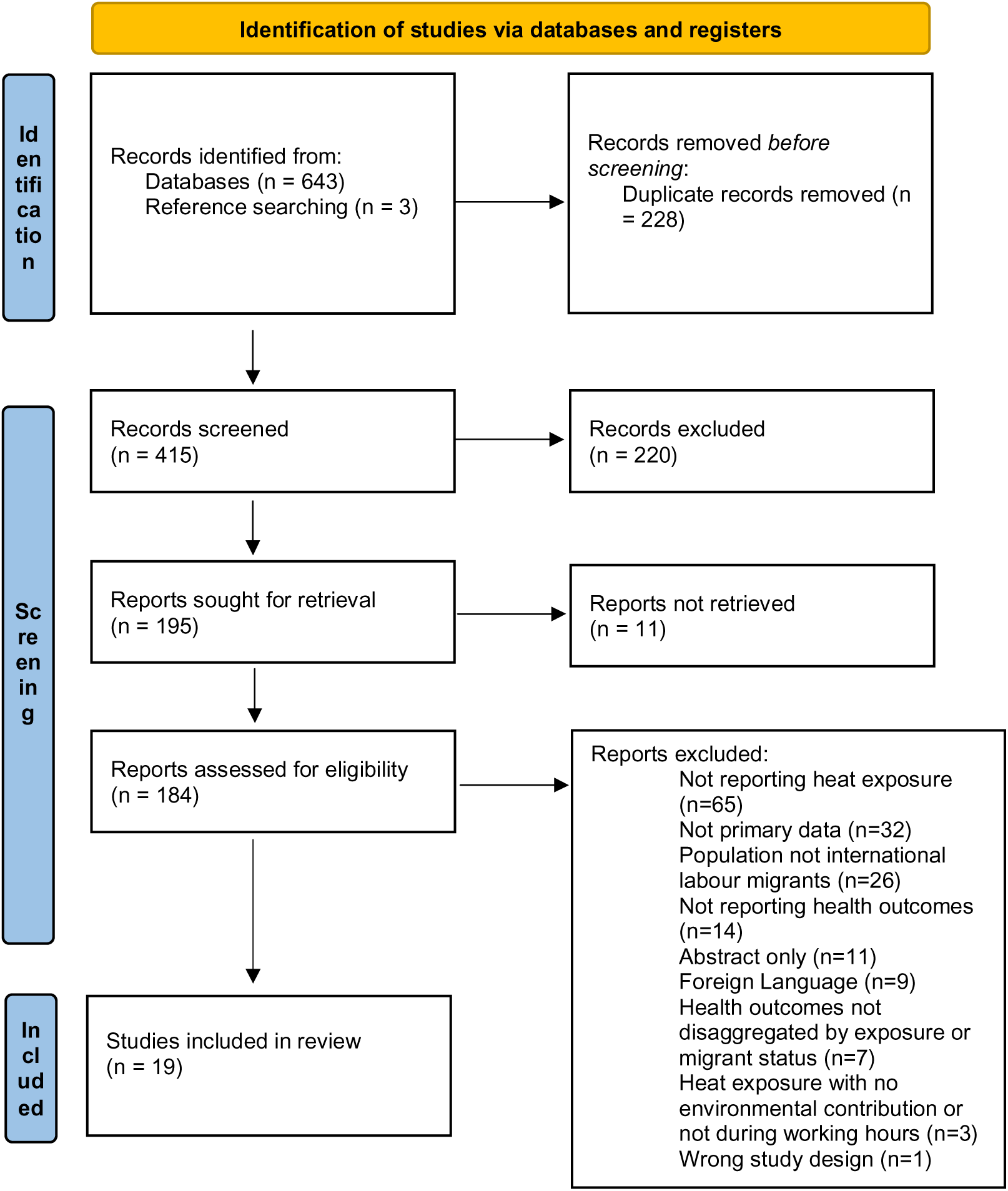
PRISMA flowchart of included studies

**Figure 2:**
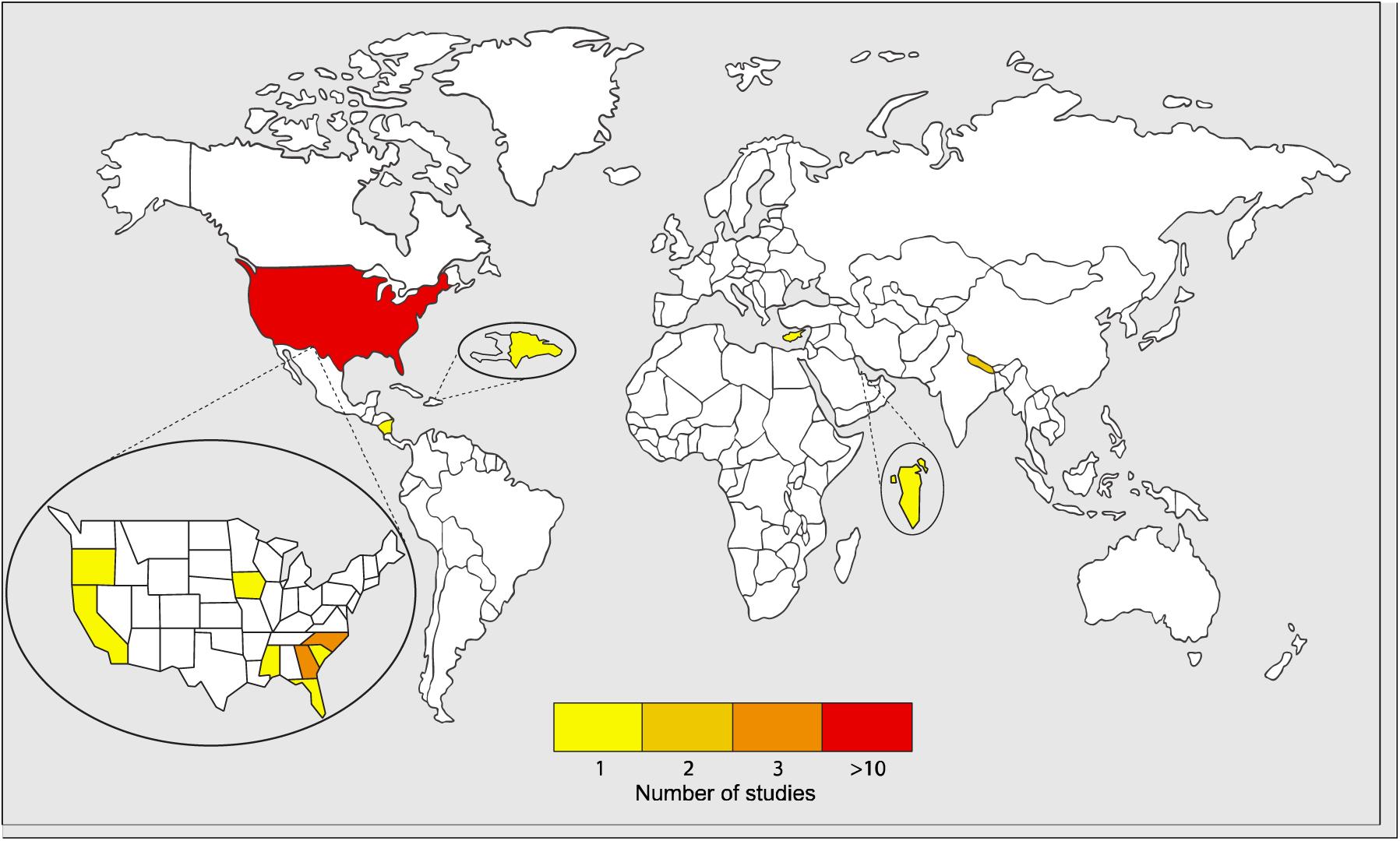
Global distribution of included studies. Figure shows that most studies are distributed in North America. Other studies are in Costa Rica (n=1) (47), The Dominican Republic (n=1)(43), Cyprus (n=1)(17), Bahrain (n=1)(34), and Nepal (n=2)(39, 49). Within North America, 3 studies were conducted in Georgia (40, 41, 45), 3 in North Carolina (37, 38, 44), and 1 in each of Florida, Iowa, California, Mississippi, South Carolina, and Oregon (33, 35, 36, 42, 46, 48).

**Table 1.** General characteristics of included studies

| Study | Study period | Location | Setting | Population | Migrant population (N) | Study focus | Quality score |
| --- | --- | --- | --- | --- | --- | --- | --- |
| **Abasilim, Friedman (33)** | 2021-2022 | Florida, USA | Vegetable farm | Migrant farmworkers | 111 | Dehydration | 5 |
| **Al-Sayyad and Hamadeh (34)** | 2008 | Bahrain | Workers' health centre | Labourers | 1111 | Climate-related health conditions | 3 |
| **Arnold, Arcury (37)** | 2017 | North Carolina, USA | The Community | Latinx child farmworkers | 30 | Heat-related illness | 3 |
| **Culp and Tonelli (35)** | 2019 | Iowa, USA | Farms | Hispanic Farmworkers | 155 | Heat-related illness | 4 |
| **Crowe, Nilsson (47)** | 2011 | Costa Rica | Sugarcane farm labour camp | Sugarcane harvesters | 91 | Heat-related symptoms | 3 |
| **Ioannou, Testa (17)** | 2016-2019 | Cyprus | Agricultural farms | Agricultural workers | 92 | Occupational heat strain risk | 5 |
| **Kearney, Phillips (38)** | 2014 | North Carolina, USA | Migrant labour camps, housing and barracks | Latino migrant farmworkers | 157 | Sun protection behaviors | 5 |
| **Keeney, Quandt (36)** | 2021 | California, USA | Not specified | Latina farmworkers | 60 | Work-life stress | 6 |
| **Luque, Bossak (46)** | 2017 | South Carolina, USA | Clinics; a migrant head start facility | Hispanic farmworkers | 29 | HRI knowledge, attitudes, perceptions and beliefs | 2 |
| **Luque, Becker (45)** | 2018 | Georgia, USA | Farmworker housing units | Hispanic farmworkers | 99 | Heat-safety knowledge, prevention, HRI risk perception | 3 |
| **Madaras, Stonington (50)** | 2019 | USA | Community health clinic | migrant farmworker | 1 | Health care, social distance, and mobility | 2 |
| **Mizelle, Larson (44)** | 2020 | North Carolina, USA | Health centre | Latino farmworkers | 30 | Fluid intake and hydration status | 6 |
| **O'Connor, Enriquez (43)** | 2017 | Dominican Republic | Mobile clinic | Dominican batey communities | 41 | Foot health | 3 |
| **Pokhrel, Ghimire (49)** | 2018 | Nepal | Hospital obstetrics and gynaecology department | Male partners of infertile couples | 86 | Infertility risk factors and semen abnormality | 5 |
| **Sharma, Koirala (39)** | 2023 | Nepal | Haemodialysis centres | Endstage renal disease patients | 95 | Environmental and occupational exposures | 2 |
| **Smith, Ferranti (40)** | 2018 | Georgia, USA | Mobile worksite clinic | Migrant farmworkers | 60 | Knowledge of HRI first aid | 6 |
| **Smith, Mac (41)** | 2019 | Georgia, USA | Hospital emergency haemodialysis services | Undocumented workers | 50 | Occupational exposures | 5 |
| **Stoklosa, Kunzler (48)** | 2020 | Mississippi, USA | Hospital emergency department | Migrant agricultural worker | 1 | Pesticide exposure, heat exhaustion, labour trafficking | 4 |
| **Wilmsen, Castro (42)** | 2019 | Oregon, USA | Forestry services network | Latino forest workers | 23 | Occupational injuries | 3 |

Regarding the social characteristics of migrant workers, ages ranged from 10 to 90 years old, of studies reporting information on sex (n=1282 participants), about 76% were male (N=973) versus 24% female (N=309). Countries of origin included India (n=865), Mexico (n=573), Nepal (n=227), Bangladesh (n=100), Nicaragua (n=91), Pakistan (n=78), Haiti (n=41), Romania (n=40), Guatemala (n=19), Vietnam (n=18), Philippines (n=12), El Salvador (n=5), Bulgaria (n=3), and Honduras (n=3) (17, 33, 34, 36–43, 45–50). Work sectors included construction (N=1113, 48%) and agriculture (crop production) (N=971, 42%), with fewer studies including migrants in services (e.g. cleaning and restaurant work) (N=30, 1%), forestry (N=23, 1%), and manufacturing (N=14, 1%).

### Heat exposure and related health outcomes

Heat exposures were described in various ways. Temperatures were reported as wet bulb globe temperature (WBGT) (17, 33, 44, 47), ambient temperature (45, 46), and heat index (40). WBGT has a range of 13.8°C (18.7 – 32.5) (17, 33), and ambient temperature 10 °C (33.3 – 43.3) (46, 48). Non-numerical descriptions of heat exposure include the words *hot* and *humid* - often together, *high temperatures*, and *occupational heat exposure* (35–37, 39, 41, 46, 48, 49).

A range of poor health outcomes related to occupational heat exposure were reported (Table 2). Heat strain was the most commonly reported (n=6) (17, 33, 35, 40, 45, 46), followed by dehydration (n=5) (33, 44, 46–48). Other reported outcomes included infertility (49), kidney disease (39, 41, 48) and compromised skin health (including dry skin and sun damage) (38, 43). One study identified a risk to pregnancy as a poor health outcome (50). Two studies also reported mental health outcomes (33, 36). Heat-related illness (HRI) was reported in three studies with no further specification or clear definition (34, 37, 42), using diagnoses of heat strain or heat exhaustion (17, 39, 47, 48), or through symptomatology (33, 35, 40, 45, 46). Five studies had sufficient data on symptoms of HRI to be collated (Figure 3) (35, 40, 45, 47, 48). The most common symptoms among migrant workers related to HRI were extreme thirst (n=90) and fever (n=80), confusion (n=60), headache (n=60), difficulty breathing (n=70), and swollen hands/feet (n=50). Additional symptoms experienced by workers were heavy sweating (n=40), skin rash (n=40), muscle cramps (n=50), tachycardia (n=30), vomiting (n=30), dizziness (n=20), extreme weakness (n=20), nausea (n=10), and pounding chest (n=10). For symptoms reported without prevalence data, a minimum of one person reporting it was assumed (33, 35, 46). Symptoms reported only once in total included heavy breathing, simultaneous hot and cold feeling, heart palpitations, dry skin, muscle spasms, stomach cramps, and weakness (35, 45, 46).

**Table 2.**
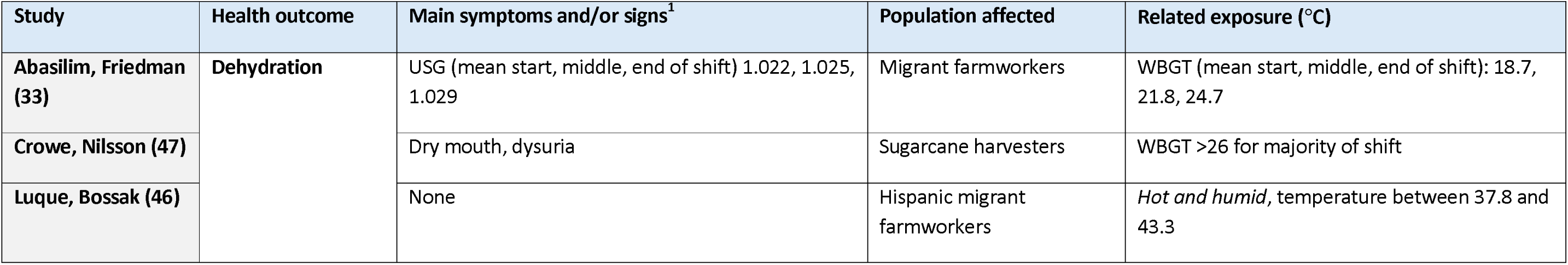
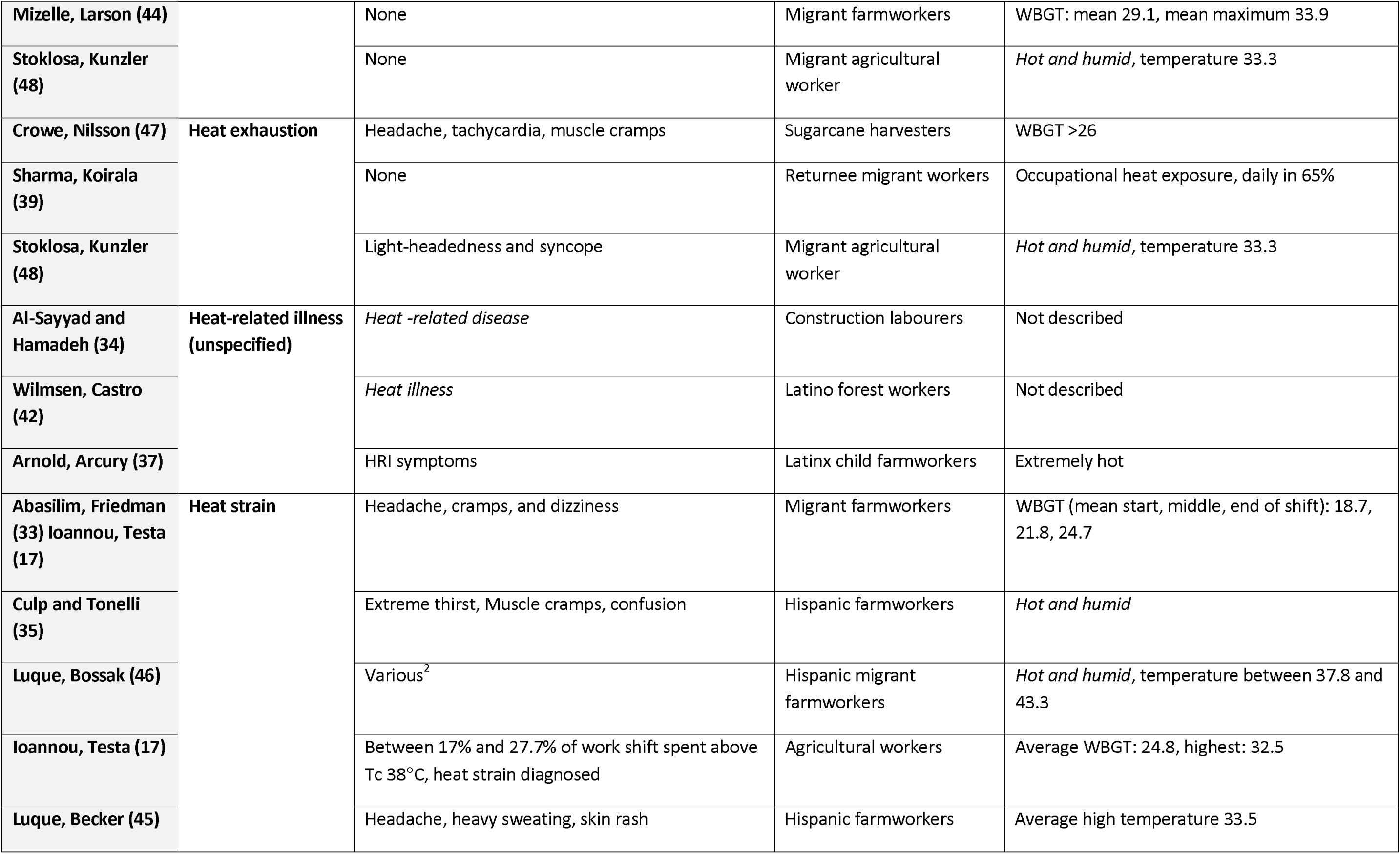
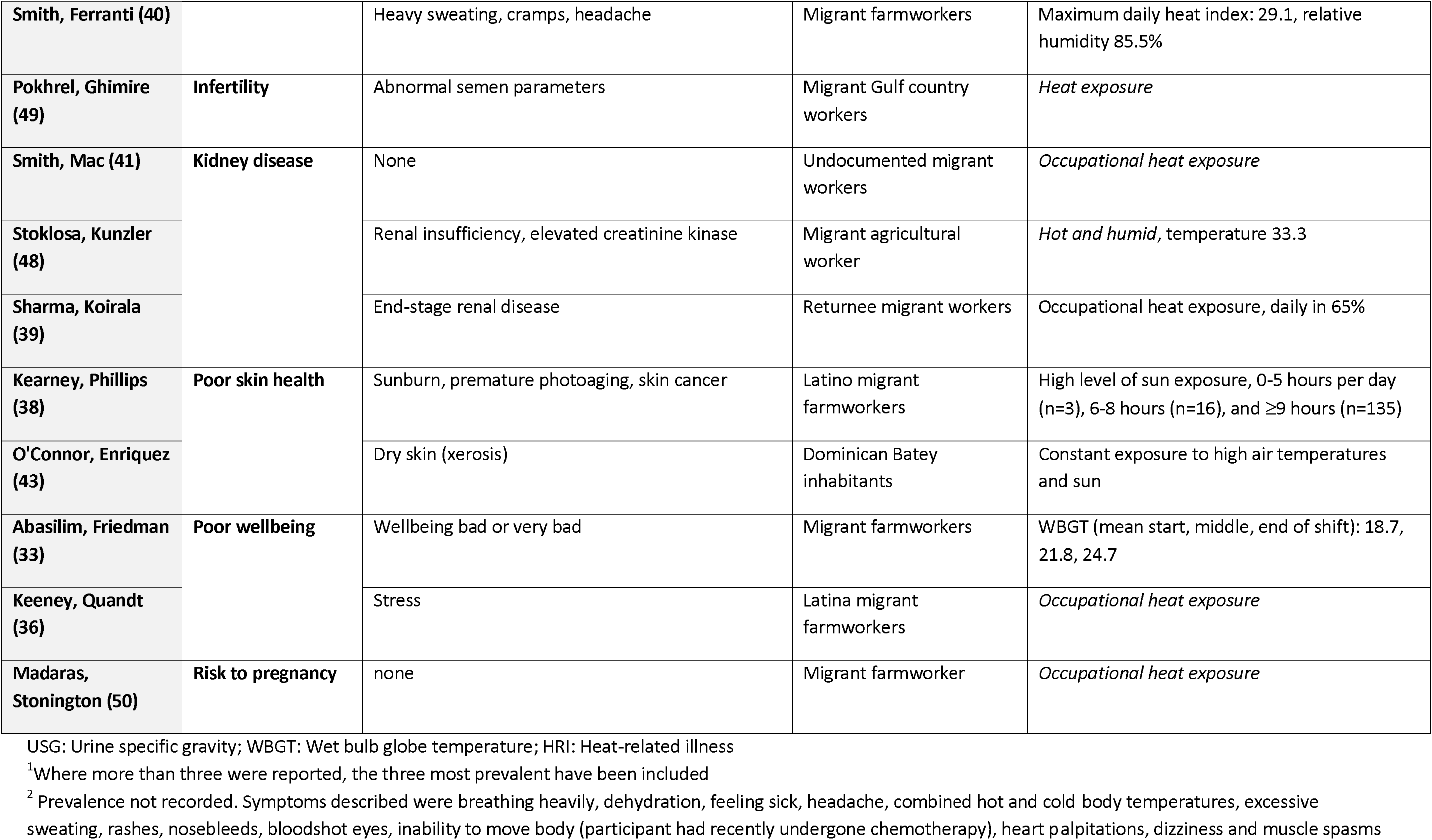
Summary of reported health outcomes in the included studies

**Figure 3.**
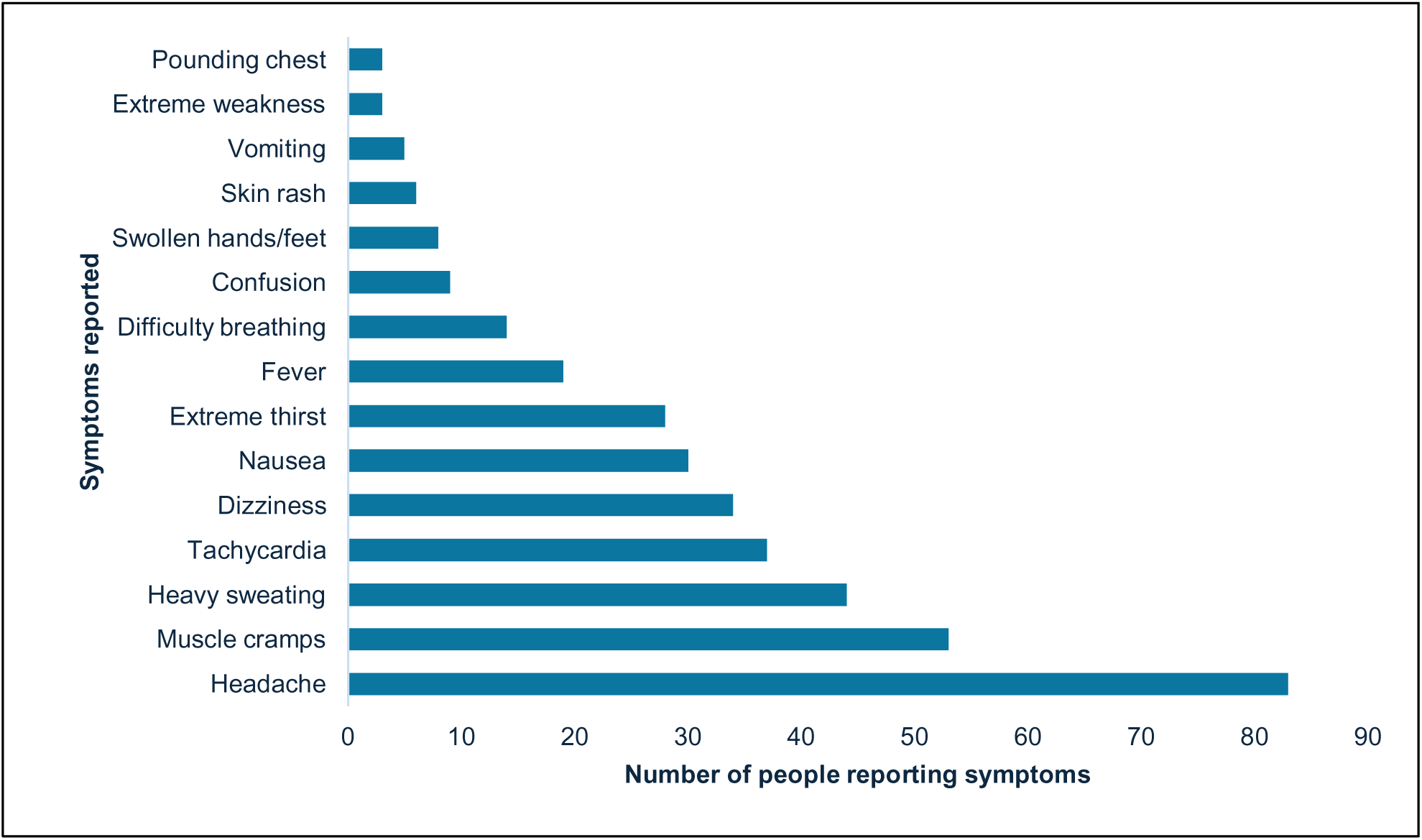
Prevalence of heat-related illness symptoms reported in included studies. Figure shows the most and less common symptoms of heat-related illness reported within 7 included studies (33, 35, 40, 45–48).

### Protective interventions and strategies to mitigate the impact of occupational heat

Twelve studies described interventions and strategies to mitigate heat-related poor health outcomes in migrant workers (Table 3). The main categories of interventions included those related to water, rest, shade, skin protection, education, work environment, healthcare services, and international guidelines. Water-related interventions, including the availability of water in the workplace, were recorded, yet challenges persisted. Where water, drinking vessels, and time to drink were provided, dehydration still worsened throughout shifts and workweeks (33). When cold water was supplied, workers reported unpleasant symptoms like muscle spasms and lung cramps, and coolers were reportedly misused to store alcohol (46). Despite adherence to NIOSH hydration standards, workers reported being inadequately hydrated, with stomach pain from bending during work further discouraging fluid intake (44). Protective measures related to rest time were reportedly met with resistance from vulnerable workers who refused to ask for breaks (35). While some workers were reported to seek shade to reduce the impact of OHS, safe shaded areas were scarce, sometimes leading to unsafe practices like resting under trucks (45). Skin protection strategies, such as wearing long-sleeved clothing, were reported to be effective in shielding workers from solar radiation (35), but in the case of headgear, the most protective style interfered with work tasks, leaving areas like the neck and face exposed when unworn (38).

**Table 3.**
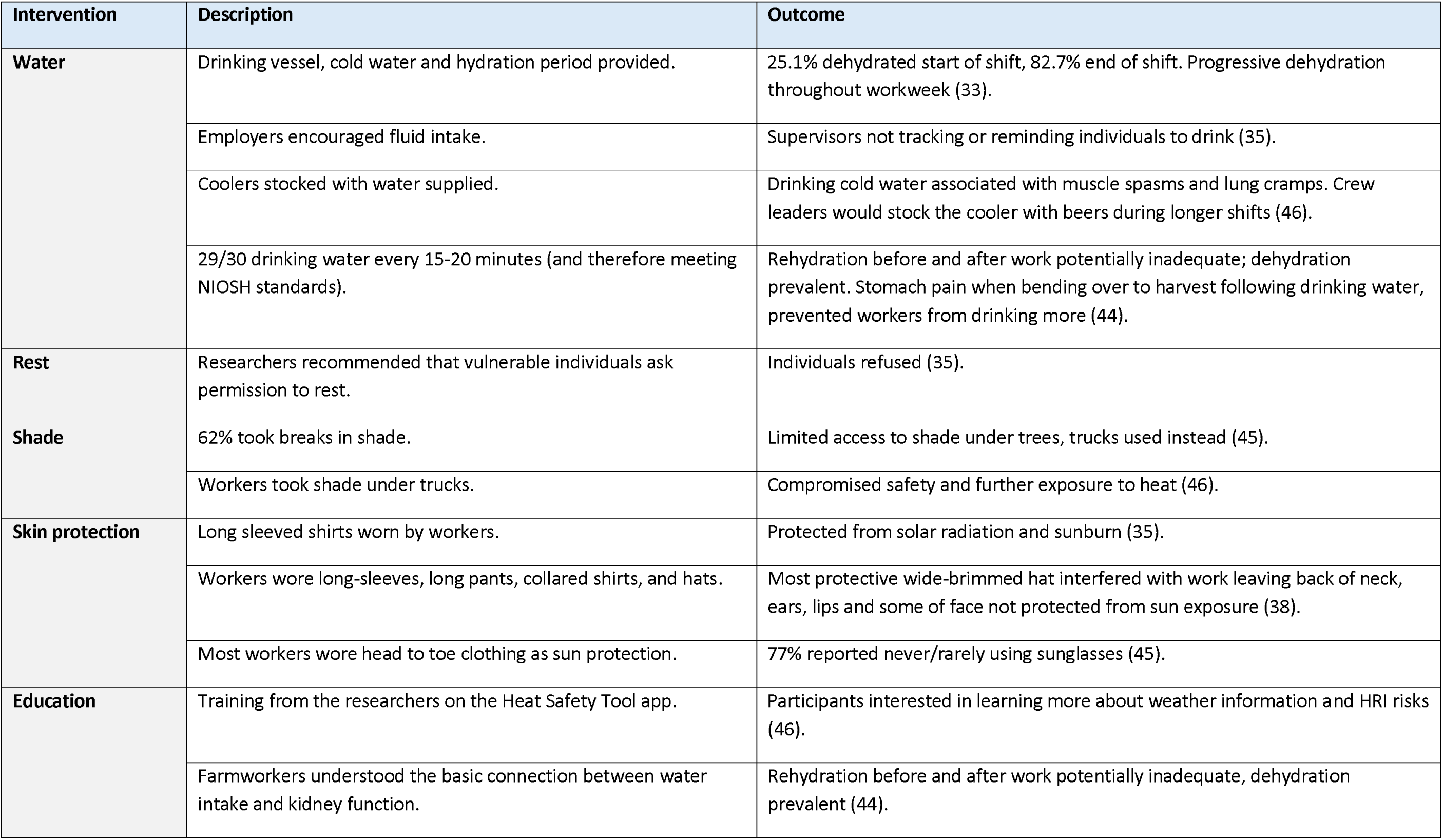
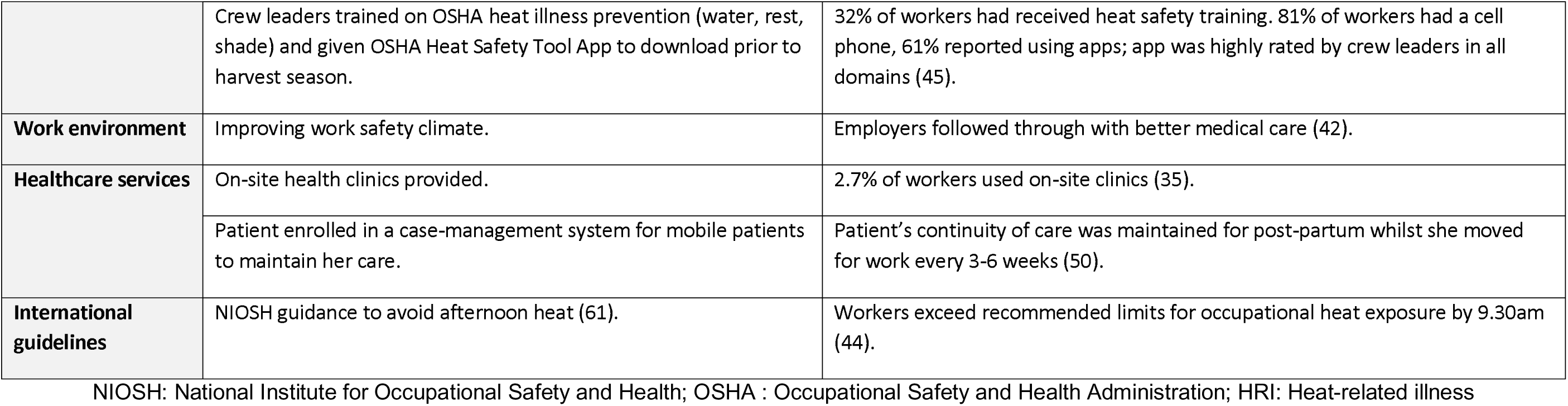
Summary of protective interventions, strategies, and their documented impact

Regarding educational interventions, workers expressed interest in learning about heat-related illness and its risks (46), however a knowledge-to-action gap remained as awareness of the links between hydration and kidney health did not translate into adequate hydration practices (44). The Occupational Safety and Health Administration (OSHA) training and the Heat Safety Tool app were well-received by crew leaders, yet only 32% of workers reported receiving heat safety training, despite 81% having access to mobile phones with which to utilise the app (45). Improvements to the working environment, such as better work-safety parameters, led to better medical care provision (42), but healthcare services like on-site clinics had low utilization amongst workers, despite higher percentages of heat-related illness symptoms (35). Language barriers further complicated access to care, as many workers lacked proficiency in the local language, limiting medical consultations and exacerbating work-life stressors (36, 42, 48) . Mobile case management systems, however, proved effective in maintaining continuity of care for a mobile worker (50). International guidelines, like NIOSH recommendations to avoid afternoon heat, were insufficient in practice, with workers exceeding recommended heat exposure limits by mid-morning (44).

## Discussion

This global systematic review identified heat-related health risks among 2322 migrant workers across six countries, with over 80% employed in construction and agriculture sectors. The study highlighted adverse health outcomes among migrant workers exposed to occupational heat, with the most commonly reported impacts being heat strain and dehydration. Other reported health issues included kidney disease, infertility, compromised skin health (such as dry skin and sun damage), mental health effects, and risks to pregnancy. A range of symptoms related to heat-related illnesses were reported, with the most common being extreme thirst and fever, confusion, headache, difficulty breathing, and swollen hands or feet. Additional symptoms experienced by workers were heavy sweating, skin rash, muscle cramps, tachycardia, vomiting, dizziness, extreme weakness, nausea, and pounding chest. Despite these significant risks, we found limited research on health-related outcomes in migrant workers, particularly in LMICs. Various protective measures have been described, including water availability, rest breaks, shade, skin protection, education, workplace improvements, healthcare services, and some level of adherence to international safety guidelines. However, significant barriers persist, such as limited access to water and hydration breaks, reluctance to take rest periods, insufficient shaded and safe areas, practical challenges with protective clothing, gaps in translating knowledge into action, underutilization of healthcare services in some cases due to language barriers, and poor implementation or inadequacy of safety guidelines.

The finding that most of those who are affected by heat stress work in construction and agriculture is in line with ILO calculations from 2019 that these two sectors would be the worst hit by reduced labour productivity resulting from heat stress (4). The geographic spread of the studies included in this review shows that there is a high concentration of reporting in HICs, particularly in the US, where workers are predominantly South American migrants, and a single study in Bahrain reporting a large population of migrant workers originating from India making it the largest group in this study (34). Migration from Mexico to the US represents the largest migration corridor globally, and data on financial remittances sent by international migrant workers show that the sources of these are nearly always sent from HICs (3). Consistent with our findings, a recent scoping review on OHS among outdoor migrant and ethnic minority workers found that most studies were conducted in the US (23). In this review, the weighted prevalence of experiencing at least one HRI symptom was estimated at 48.8%, while 27.7% experienced at least three symptoms with higher prevalence rates in studies outside the US (23).

Indeed, trends in global exposure to extreme heat in relation to disease burden show that LMIC populations experienced a higher risk of exposure to extreme heat in 2010-2019, and a subsequent greater health loss than HICs (51). Thus, research informed policy around heat and health in these countries is required to fill the knowledge gap regarding international migrant workers in LMICs. Occupational heat exposure also affects migrant workers upon their return to their home countries, for example in the case of Nepal as found in this review (39, 49). Other countries in the Asian sub-continent also receive a large amount of international remittances from emigrated workers (3), indicating that protective interventions to mitigate occupational heat exposure could also be targeted towards international migrant workers upon their return to their home countries.

Our findings also indicate that analyses to compare evidence on heat-related illness had several limitations. Difficulties categorising HRI arose due to the inconsistent terminology used in the included studies. Diagnoses of heat-strain, heat-related disease, heat exhaustion, and heat illness were all used to describe experiences of HRI (17, 34, 39, 42, 47, 48). To understand the burden of heat-illness in this vulnerable population, more specific classification is required. Wight, Wimbush (52) explain the process of public health intervention development, the first step being the clarification of the problem, emphasising the need for clear definitions of the health issue and its cause. Reporting of heat exposure is not standardised in the included literature and is often poorly defined. Temperatures were measured using different methods, with only few studies using WBGT - the method recommended by OSHA for monitoring workplace heat levels (17, 33, 44, 47, 53). Increased consistency in exposure and outcome reporting is needed for the creation of effective, targeted interventions for OHS and to enable cross-study comparisons. This issue has been reported by other researchers on similar topics (23), limiting such comparability.

The findings from the systematic review reveal progress and gaps in interventions and measures aimed at protecting migrant workers from heat exposure. While the OSHA-recommended framework of water, rest, and shade has informed many measures in this review, its implementation often falls short, as seen in cases where inadequate shade or misuse of resources, such as coolers stocked with alcoholic drinks, exacerbated risks (45, 46). Educational interventions have shown promise in improving safety knowledge (54), but their reach remains limited, with training and safety information sometimes failing to reach workers (35, 45). Farmworkers’ expressed desire for more information on heat risks (46). underscores the need for worker-centered approaches, including direct education and involvement in designing safety measures. This demonstrates how systemic barriers such as weak enforcement of regulations and lack of employer accountability must be addressed to ensure effective and sustainable protections.

Shortcomings in hydration practices and healthcare accessibility for migrant workers exposed to heat are also highlighted by this review. NIOSH standards recommending water intake every 15-20 minutes were insufficient to prevent dehydration, which worsened despite the provision of water, drinking vessels, and designated breaks (33, 44). Regular water intake alone failed to counteract dehydration, a risk factor for AKI (44, 55), which is also exacerbated by hyperthermia and physical labor (56). Electrolyte drinks and oral rehydration solution (ORS) have been identified as more effective in replenishing lost fluids and electrolytes, with ORS showing superior fluid retention during exertion (57, 58). These findings suggest a need to investigate ORS as a targeted intervention for dehydration and AKI prevention in this population. Additionally, despite the availability of onsite clinics, usage remained low (35), and language barriers increased difficulties to healthcare access in other studies (36, 42, 48). Language barriers, low income, lack of health insurance, and preferences for self-medication have been identified as key factors limiting healthcare access and utilization in other populations of migrant workers (59). Further research across diverse settings is needed to explore barriers to healthcare access in work environments, and develop strategies to improve service utilization among international migrant workers.

We found that personal protective equipment (PPE) designed to protect against sun exposure went unused because they interfered with workers’ tasks (38), and suggestions to take breaks were refused by vulnerable workers (35). This example illustrates that interventions lacking suitability for the target population might to see low uptake and fail to achieve desired outcomes. In contrast, a backpack hydration system developed with farmworkers’ mobility in mind was widely accepted for use amongst migrant workers and resulted in increased water intake (60). Similarly, a recent review showed that although regular breaks are allowed, some migrant and ethnic minority workers forgo them due to barriers such as the desire to earn more or for fear of losing their jobs (23). This demonstrates the value of involving workers in the design process to ensure practicality and acceptability of interventions and to address barriers to the uptake of such measures.

In light of these findings, recommendations are focused on strengthening occupational heat exposure research amongst migrant workers in LMICs. Specific investigations should address barriers to occupational health services amongst migrant workers at risk of heat-related illness and the efficacy of ORS for improving hydration in working migrants. Going forward, heat illness prevention strategies should be enhanced using worker targeted education and increased accessibility to weather monitoring and heat-safety information systems. Strategies to develop additional heat-protective interventions should prioritise participatory approaches engaging migrant workers (Figure 4).

**Figure 4.**
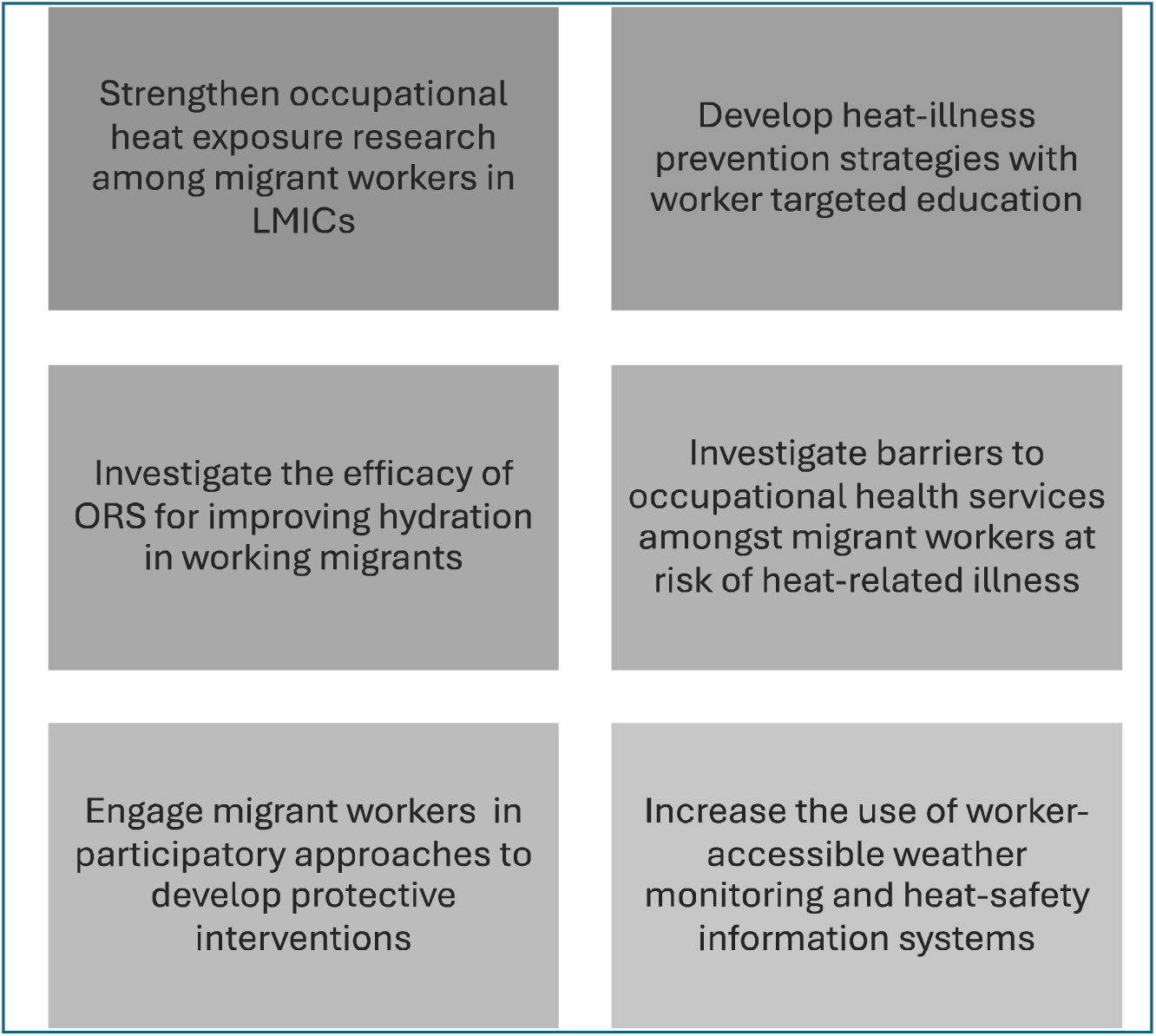
Recommendations for future research and policy

This review has some limitations. One is the exclusion of articles in languages other than English. As this study was aiming to gain a global perspective on the impact of heat exposure on migrant workers, excluding those in a foreign language may have missed important findings in migrant populations affected by occupational heat exposure elsewhere, resulting in a distorted view of study reporting on this topic. Another limitation of this study is the large number of low-quality studies. Broad inclusion criteria regarding study quality were required to ensure all available data on this topic was investigated. The inclusion of studies assessed as low quality was deemed to be important due to the scoping nature of this review in a new and expanding field. For the same reason, this review does not identify health outcomes for which the confirmed cause is exposure to high environmental temperatures, only that which may be associated with such an exposure.

## Conclusions

This systematic review has identified a population of migrant workers worldwide whose health was adversely affected by occupational heat exposure. Interventions being implemented to mitigate against adverse health effects related to occupational heat exposure were identified. Unsuccessful outcomes from these interventions, as well as worker characteristics and behaviours delineate the specific factors mediating poor health outcomes in migrant workers. Recommendations following these observations are that interventions must be migrant focused, and interventions should be combined with heat-safety education of all workers and employers, which will improve their success and sustainability. These findings should inform the creation of policy and guidelines protecting migrant workers’ health. This review has found that, despite guidelines surrounding temperature limits, water drinking standards, and adaptive behaviours, migrant workers are still burdened by occupational heat exposure with associated health effects as severe as requiring hospitalisation. With the effects of global warming only intensifying, correction of these shortfalls in protecting the health of migrant workers is rapidly required.

## Ethics approval and consent to participate

Not applicable.

## Availability of data and material

Data are available on reasonable request from the corresponding author.

## Author’s contributions

LH and KL generated the protocol with input from SH. LH conducted the database search. LH and MJ performed title/abstract screening and full-text review. Data extraction was done by LH and SW. Risk of bias assessments were conducted by LH and MJ. LH led the data analysis with input from SH and SW. The first draft of the manuscript was written by LH and OB, with contributions from SH and all other authors. SH supervised the work, and SH and LH accessed and verified the data. Additional contributions were made by BF, AZ, IE, TL, AF, DT, and CZ on behalf of the Consortium for Migrant Worker Health.

## Data Availability

All data produced in the present study are available upon reasonable request to the authors

