## Supplementary Data for "International Migrant Workers, Heat Exposure, and Climate Change: A Systematic Review of Health Risks and Protective Interventions"

**Table S1. Search terms**

| **Search concept 1: International migrant workers** | |
| --- | --- |
| 1 | ((foreign* OR non-native* OR non-national* OR migrant* OR refugee* OR undocumented OR expat* OR traffick* OR immigrant*)adj3(work* OR labo?r OR labo?rer OR job OR staff OR occupation* OR employ* OR forestr* OR plantation* OR manufactur* OR construction OR farm* OR brick kiln OR factory OR sugarcane OR fisher* OR mining)) |
| 2 | economic adj2 (migrant OR immigrant) |
|  | **1 OR 2** |

**AND**

| **Search concept 2: Health outcomes** | |
| --- | --- |
| 1 | health OR injur* OR disease* OR hazard* OR exposure* OR accident* OR hygiene OR safety OR trauma* OR fatal* OR harm* OR death* OR ill OR illness* OR sick* OR syndrome* OR wound* OR fatigue OR risk* OR disabilit* OR morbidit* or mortalit* OR infect* OR disorder* OR condition OR pain OR pains OR sore* OR ache* OR unwell OR hospital* OR vulnerabilit* OR strain* OR stress* OR danger* OR well?being |
| 2 | respirat* OR musculoskeletal OR cardiovascular OR cancer* OR hypertensi* OR dermatitis OR allerg* OR fall* OR drown* OR exhaust* OR broken bone* OR toxic* OR burn* OR parasite* OR pesticide* OR insecticide* OR hydration OR asthma OR bronchitis OR pulmonary OR fatigue OR nephropathy* OR renal function OR urolithiasis OR kidney* OR ckd* OR aki OR anxi* OR depress* OR psychiatr* OR psycho*OR suicid* OR distress* |
| 3 | Dehydrat* OR exhaust* OR fatigue* OR syncope OR edema OR oedema OR HRI OR sunburn* OR cramp* OR rash OR miliaria OR stroke OR hyponatremia OR hypernatremia OR rhabdomyolysis |
|  | **1 OR 2 OR 3** |

**AND**

| **Search concept 3: Heat exposure** | |
| --- | --- |
| 1 | heat* OR warm* OR hot OR dry OR summer OR temperatures OR wbgt OR humid* OR arid OR ultraviolet OR UV OR drought OR thermal |
| 2 | sweat* OR hypertherm* OR strain OR electrolyte* OR tolerance |
| 3 | ((climate OR climatic)adj2(change OR event OR disaster OR crisis OR variation OR variabilit*)) OR extreme weather |
|  | **1 OR 2 OR 3** |
